# Occupational hazards and risks among the women in fisher communities in Cox’s Bazar and Chattogram

**DOI:** 10.1101/2024.01.08.24300962

**Authors:** Charls Erik Halder, Partha Pratim Das, S.M. Tareq Rahman, Liton Chandra Bhoumick, Hamim Tassdik, Md Abeed Hasan, Sourav Nath Mithun

## Abstract

**Background:** Women in the fisher communities in coastal regions of Bangladesh are engaged in a wide range of fishery activities including fish sorting, grading, cutting, dry fish processing, transporting and selling. However, there is limited evidence available on the occupational hazards and risks experienced by them.

**Method:** The study was conducted among fishing colonies in Cox’s Bazar and Chattogram districts in Bangladesh. This was a cross-sectional study comprised of both qualitative and quantitative approaches. Data were primarily collected through eight focus group discussions (FGD) and a quantitative survey of a sample of 207 women working in fisheries sector.

**Findings:** The study found a high occurrence of occupational hazards, health risks and disease conditions and limited availability of preventive measures among the women in the fisher communities in the coastal Bangladesh. Occupational hazards include physical safety hazards, like slippery surface, fish cutting instruments, fish sting or bite and contact with fishes; physical hazards, like prolonged sun exposure and noise; chemical hazards like pesticides, salt and salt water; ergonomic hazards, like prolonged sitting or standing in uncomfortable posture and heavy weight lifting; and biological hazards, like inadequate provision of sanitary latrine or hand washing soap at workplace. The study also found occupational risks resulting from the hazards including injuries (87.44%), musculoskeletal conditions (69.08%), skin diseases/conditions (56.52%), eye complaints (33.82%), severe respiratory distress (24.15%) and high incidence of self-reported communicable diseases. Majority of the women did not use personal protection equipment at their workplace (78.26%) and have a first aid kit at their workplace (93.72%).

**Conclusion:** This study highlights the critical occupational health and safety challenges faced by the women in the fisheries sector. A comprehensive multisectoral strategy needs to be undertaken to mitigate the occupational hazards and prevent associated diseases among the women in fisher communities promoting their health and wellbeing.

## Background

Globally, fisheries and aquaculture sector produce more than two hundred million tonnes of fish, aquatic animals and algae critically contributing to global food security and nutrition. ^1^ This industry employs over 58.5 million people worldwide, with women making up 21% of those workers. ^2^ In Bangladesh, around 17 million people depend on fishing, fish farming, fish handling, and fish processing for their sustenance.^3^ Marine fishery is vital component in Bangladesh’s fishing industry because of the country’s extensive coastline and marine zone.

Around 30% of the women in rural and coastal areas of Bangladesh are engaged in fisheries sector.^4^ Women in fisher communities in Bangladesh have a twofold dilemma since patriarchal societal norms limit their activities to the home, but rising poverty and landlessness also force them to look for wage work. Thereby, they are to play both role, fishing activities as well as traditional household chores. An earlier baseline study in Cox’s Bazar and Chattogram revealed that women in the fisher communities were engaged in a wide range of fisheries activities including fish sorting, grading, cutting, dry fish processing, dry fish paste (*Nappi*) processing, daily laboring in fish farms, transporting, selling fish or fish products, net making, ice processing, fish basket making, and collecting minnow and seashells from the sea shores.^5^ Still, traditionally, fishing and related activities in Bangladesh are considered to be the occupation of men in Bangladesh, particularly in coastal regions, and women’s contributions to this sector are grossly neglected.

Globally, people engaged in fishery sector encounter numerous occupational hazards, including the risk of boat capsizing, slippery surface, unsafe handling of heavy weight, fish bites and stings, snake and insect bites and exposure to physical hazards, like sun, salt water and noise, which may result in various illness and deteriorated heath conditions, including injuries, fatalities, skin, respiratory and auditory diseases.^6–8^, According to the World Health Organisation (WHO) and the International Labour Organisation (ILO), globally, almost 2 million individuals died as a result of occupational hazards in 2016.^9^

The Committee on Fisheries (COFI) declaration for sustainable fisheries and acuaculture emphasized to enhance women’s full access to and equal opportunities in the fisheries sector.^10^ To achieve this, occupational safety and health of the women engaged in the fisheries should be a key consideration. There are several studies available both at country and global level on the occupational hazards encountered by the men in the fisheries sector. However, only a few studies^11–12^ available that identified occupational hazards and risk factors of women engaged in fishery activities. In Bangladesh, there is no study available on the occupational hazards of women working in fisheries, except, a qualitative study^13^ found that women engaged in fishing with their children possesses the risk of drowning since the traditional cloths are not flexible enough for free-movement in the water.

Fishery is one of the oldest and most prevalent occupations in Cox’s bazar and Chattogram coastal communities. Fishery engages a wide range of occupational activities, including production, harvesting, processing, handling, storage and transportation. Traditionally, men and women have different roles, while men are mostly engaged in harvesting, women are mostly engaged in fish processing. Therefore, women may experience distinctive hazards and risks in their occupation, which is vital to reveal to ensure a safe work environment for them. Moreover, it is also essential to understand the measures in place to prevent occupational hazards and risks, such as personal safety training and personal protection equipment, for proper planning of preventive strategies. The study was conducted to identify the different occupational hazards that women in fisher communities in Chattogram and Cox’s Bazar were exposed to, the occupational risks that ensued from those hazards, and the extent of health and safety measures that were in place to prevent occupational health hazards.

## Methodology

### Study Setting

The study was conducted among fishing communities in Cox’s Bazar and Chattogram districts in Bangladesh. Table 1 shows the detailed breakdown of the study location.

**Table 1.**
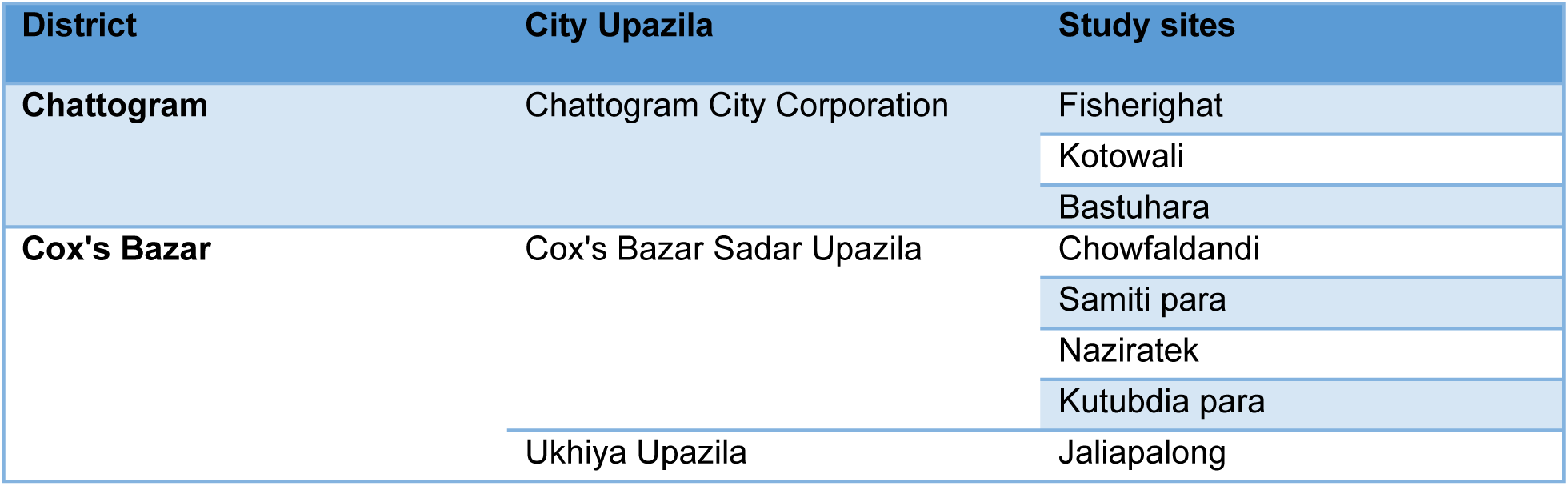
Study location.

The study sites in Cox’s Bazar are located by the Bay of Bengal and inhabited by thousands of fisherfolk communities. Livelihood in these areas is mostly tied to fishing from the sea and related fishery activities. Naziratek-Samitipara area, the largest dry fish processing zone of the country, accommodates hundreds of dry fish processing trades. Chowfaldandi area is also inhabited by indigenous Rakhaine community. A traditional delicacy of the indigenous people, powdered dry fish (also known as *nappi* locally), is prepared here and exported to the indigenous populations in various districts of Bangladesh. Study sites in Chattogram are located by the river Karnaphulli, where fisher colonies are engaged in fishing and fish processing.

### Study design

This was a cross-sectional study comprised of both qualitative and quantitative approaches aimed to explore the occupational health hazards and risks of women in the fisher communities in Chattogram and Cox’s Bazar. The study had an exploratory sequential design – qualitative data collection and analysis was done in first place to understand the occupational setting, hazards and risks and helped in development of data collection tool for subsequent quantitative survey. The recruitment period of the study was from 29^th^ August 2023 to 30^th^ October 2023.

The qualitative study comprised of six focus group discussions (FGD), two at Chattogram, two at Cox’s Bazar Sadar and one a Ukhiya Upazila. This was conducted to get in in-depth understanding of a) socio-economic situation of the women in fisherfolk, b) potential physical safety, physical, chemical, biological and ergonomic hazards experienced by them, c) potential outcomes of the hazards to different human system and d) existing situation of health and safety measures at workplace. A total of 64 women from fisherfolk participated in the FGDs, each participated by five to eight individuals. A semi-structured guiding questionnaire was used for guiding the discussion, which was developed based on previous studies. ^7,8,11,12^ The guiding questionnaire was piloted among a group of 5 fisherfolk women to ensure its validity and reliability. The principal investigator and 5 trained research assistants facilitated the FGDs. At each FGD, one research assistant was assigned as the note taker and to ensure proper audio-recording of the discussion. An interpreter fluent in local Chittagongian language was engaged at the FGDs to eliminate the linguistic barriers. Average duration of each FGD was around 90 minutes.

The quantitative component of the study included survey of women working in fisheries sector using a structured questionnaire. The questionnaire form was deployed through the Kobo toolbox for data collection. The questionnaire was developed in line with the research objectives and potential occupational health hazards and illnesses explored through the qualitative study as well as the health and safety measures recommended by the Convention 188 of the International Labor Organization (ILO).^6^ The questionnaire comprised questions in relation to a) socio-demographic profile of the participants, b) exposure to physical safety, physical, chemical, biological and ergonomic hazards, c) potential outcomes of the occupational hazards to different human systems and d) status and practice of different health and safety measures. The questionnaire was initially piloted with 10 participants, and adjustments was made as needed in the final version based on the results of the testing. 4 enumerators, who were fluent in local language, under the supervision of the research assistants were responsible for the quantitative data collection using the Kobo toolbox form.

The research assistants and enumerators were adequately trained on the research objectives and scopes, data collection instrument and procedures. The team was also oriented on the obtaining informed written consent and maintaining proper privacy and confidentiality.

### Study population, sampling and sample size

The study was implemented in 8 selected sites in Cox’s Bazar and Chattogram, which accommodates around 7,000 fisherfolk families. Study sites were purposively selected where diversified fishery activities are prevalent. To determine the appropriate sample size for the quantitative study, the researchers used a calculated occupational injury rate of 85% from a similar setting^14^ as a reference point. Cochran’s formula was applied to calculate the sample size required for a 95% confidence level and a 5% sampling error. Initially, this calculation yielded a sample size of 191. However, the researchers decided to adjust the final sample size to 207 participants considering the potentiality of unresponsiveness. Participants were selected using the simple random sampling from a local list of fisher families For qualitative study, number of each focus group discussion was between 6 to 12 as recommended by empirical studies,^15–17^ making a total of 48 participants, all were women engaged in fisheries activities. Participants were purposively selected with the assistance of local volunteers.

### Data analysis

The qualitative data, i.e. FGD transcripts, were transcribed verbatim and manually analyzed using a thematic analysis approach. This involved identifying recurring themes, patterns, and insights from the qualitative data, contributing to a richer understanding of the occupational health hazards and their impact faced by women in the fisherfolk communities. Descriptive analysis of the data collected through the structured survey questionnaire was carried out using the built-in analysis tools in Kobo Toolbox and SPSS (Statistical Package for the Social Sciences).

### Ethnical consideration

The Institutional Review Board (IRB) and Ethics Review Committee (ERC) of North South University in Bangladesh approved the research protocol (2023/OR-NSU/IRB/0810). All participants willingly gave written informed consent, primarily through thumbprints, and these consent documents were securely stored. The study adhered to the "no-harm" principle, ensuring no legal risks for participants. Each step was conducted in accordance with the venerable Helsinki Declaration (1964) and its 2013 revision.

## Results

### Demographic profile of the participants

The study engaged 207 women working in the fisheries sector in Cox’s Bazar and Chattogram. Table 2 summarized the demographic profile of the participants. Nearly half of the participants (43.48%) were aged between 31 to 40 years. More than 20% of the participants were belonging to 18 to 30 years, and a similar percentage to 40 to 50 years. Majority of the participants could not sign (64.25%) and 18.36% could only sign. 10.63% studied upto class 5 and only a few studied upto class 9 (5.80%) or passed secondary school certificate (SSC) (0.97%). No one was found to be studied beyond the level of SSC. Ethnicity of the majority of the participants were Bengali (94.20%), 5.80% were tribal belonging to Rakhaine community.

**Table 2.**
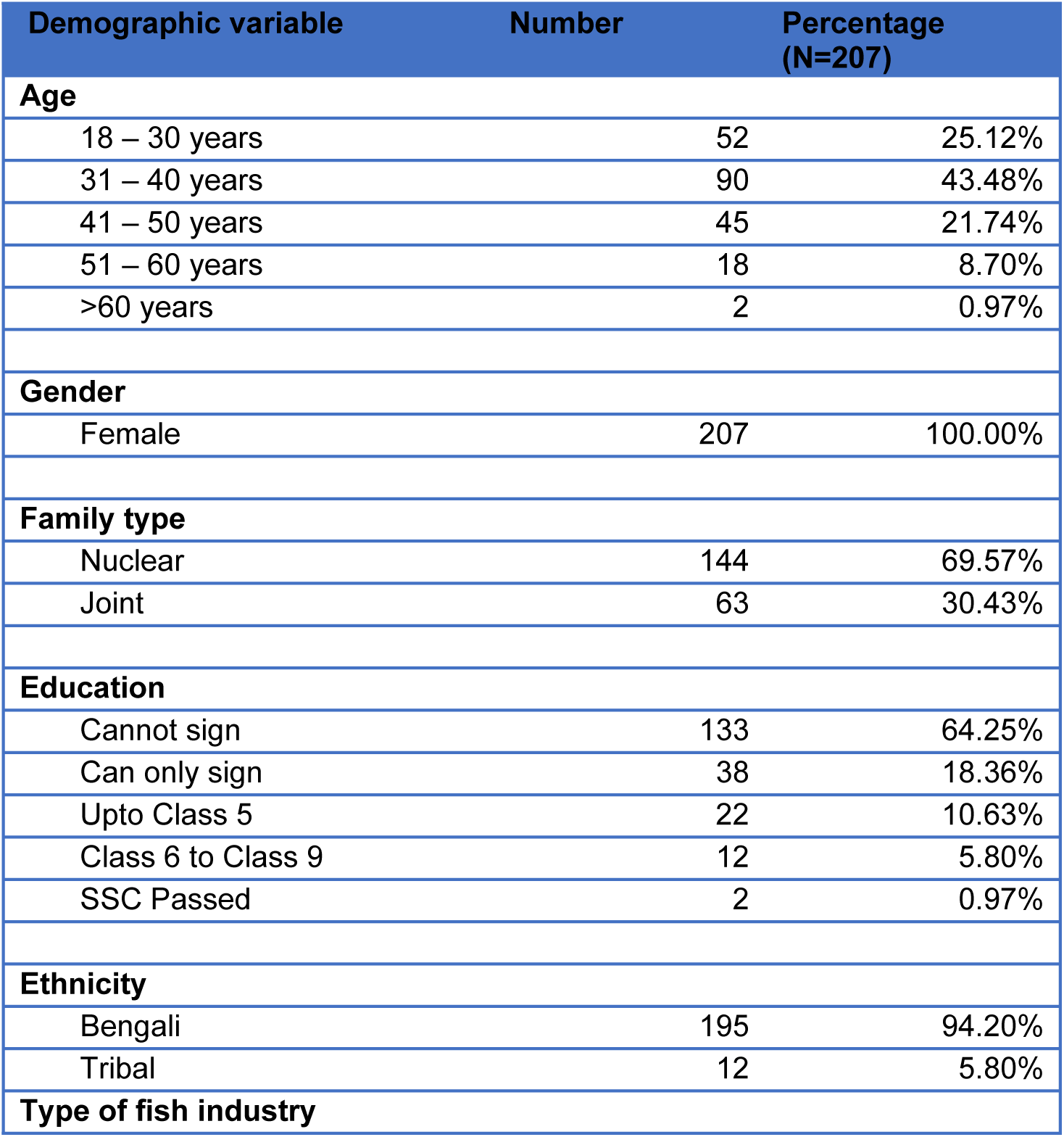
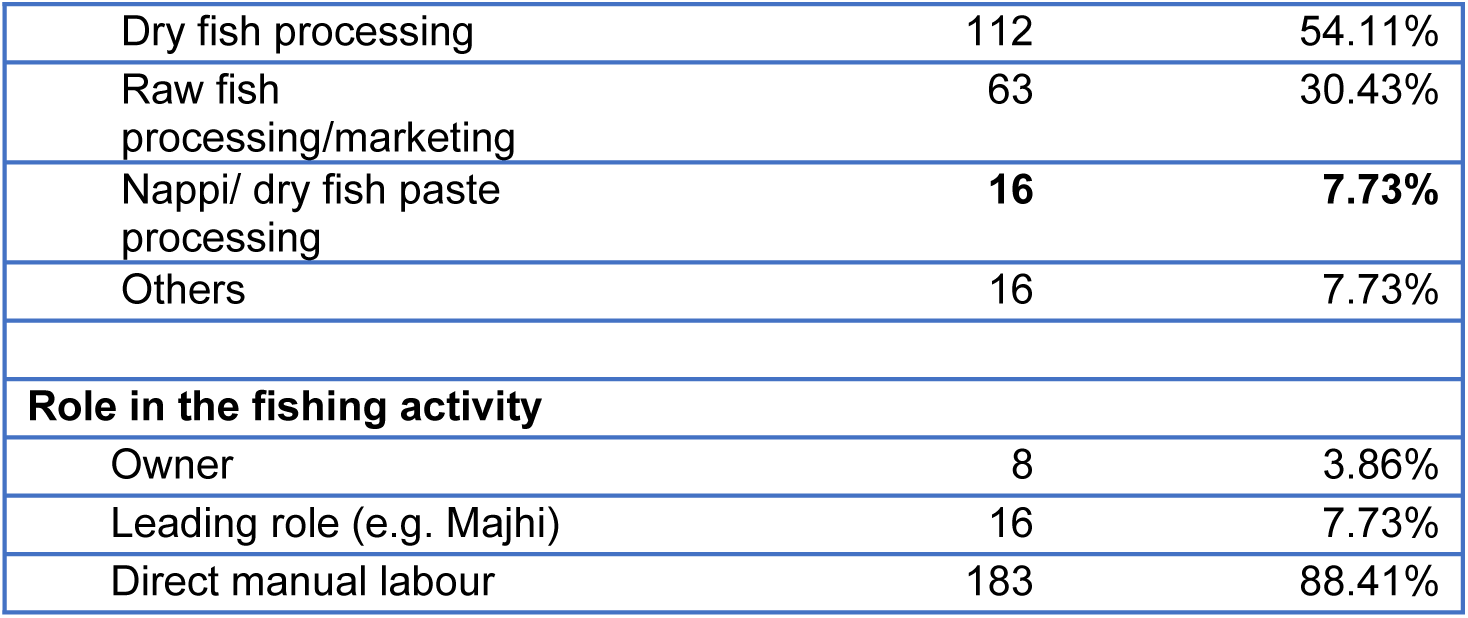
Socio-economic and demographic variables of the participants.

| Demographic variable | Number | Percentage (N=207) |
| --- | --- | --- |
| **Age** |  |  |
| 18 – 30 years | 52 | 25.12% |
| 31 – 40 years | 90 | 43.48% |
| 41 – 50 years | 45 | 21.74% |
| 51 – 60 years | 18 | 8.70% |
| >60 years | 2 | 0.97% |
| **Gender** |  |  |
| Female | 207 | 100.00% |
| **Family type** |  |  |
| Nuclear | 144 | 69.57% |
| Joint | 63 | 30.43% |
| **Education** |  |  |
| Cannot sign | 133 | 64.25% |
| Can only sign | 38 | 18.36% |
| Upto Class 5 | 22 | 10.63% |
| Class 6 to Class 9 | 12 | 5.80% |
| SSC Passed | 2 | 0.97% |
| **Ethnicity** |  |  |
| Bengali | 195 | 94.20% |
| Tribal | 12 | 5.80% |
| **Type of fish industry** |  |  |
| Dry fish processing | 112 | 54.11% |
| Raw fish processing/marketing | 63 | 30.43% |
| Nappi/ dry fish paste processing | **16** | **7.73%** |
| Others | 16 | 7.73% |
| **Role in the fishing activity** |  |  |
| Owner | 8 | 3.86% |
| Leading role (e.g. Majhi) | 16 | 7.73% |
| Direct manual labour | 183 | 88.41% |

More than half of the women (54.11%) were engaged in dry fish processing farms, either commercial or personal. Around one-third (30.43%) were engaged in raw fish processing and marketing. 7.73% of the women were engaged in dry fish paste (nappi) processing and rest were engaged in other fishery activities such as minnow or seashell collection. When we asked about day-to-day activities of the women in the fisher folks, we found that they were engaged in multiple fishery activities. Three fourth of the women (73.91%) were engaged in fish sorting and grading. Around one-third of the women engage themselves in fish cutting or removing scales (34.3%), dry fish processing (32.85%) and minnow collection (32.37%) from seashores. More than quarter of the respondents were engaged as day laborer in fishing farms (28.02%) and in seashell collection (25.12%). 17.87% of the women were found engaged in dry fish paste (*nappi*) processing. A few of the women were also found engaged in other fishery activities, such as supporting direct fishing (8.70%), selling fish or fish products (7.73%), basket production (6.28%), shopkeeping (5.31%), fishing net making or repairing (2.42%), leading fishery activities (1.93%) and transporting fish or fish products (1.93%). Majority of the women dedicated direct manual labour (88.41%) in their fishing industry, while only a few were owner (3.86%) or played leading role (Majhi) (7.73%) in the fishing industry.

### Occupational Hazards

The study identified a wide range of physical safety, physical, chemical, biological and ergonomic hazards exposed by the women working in the fisheries sector. Table 3 summarized the hazards encountered.

**Table 3.** Occupational hazards for women working in fishing industry.

| **Physical safety hazards** |  |  |
| --- | --- | --- |
| Slippery surface | 129 | 62% |
| Fishing or minnow collecting instruments | 90 | 44.4% |
| Fish cutting instruments | 79 | 38% |
| Fish sting/fish bite | 120 | 58% |
| Contact with fish | 102 | 49% |
| Sharp stone/corals or seashells | 5 | 2% |
| Basket making | 2 | 1% |
| Grinding machine of Nappi | 2 | 1% |
| Drifting away | 2 | 1% |
| **Physical hazards** |  |  |
| Sun exposure (>4 hours) | 205 | 99% |
| Noise | 65 | 31.4% |
| **Chemical hazards** |  |  |
| Chemical compounds | 89 | 42.99% |
| Salt water | 191 | 92.27% |
| Salt | 167 | 81% |
| **Ergonomic** |  |  |
| Uncomfortable body posture | 188 | 91% |
| Prolonged sitting in an uncomfortable posture | 174 | 84% |
| Prolonged standing | 103 | 50% |
| Heavy weightlifting | 108 | 52% |
| Repeated pulling/troughing or hanging | 5 | 2% |
| **Biological** |  |  |
| No sanitary latrine at workplace | 150 | 72.46% |
| No facility for washing hand with soap at workplace | 162 | 78.26% |
| Working longtime wearing wet clothes | 155 | 74.88% |

#### Physical safety hazards

The participants identified a range of physical safety hazards that caused them injuries. 62% of the participants identified slippery surface at their workplace as a hazard which causes accidental falls and injuries. 44.4% of the participants reported fishing or minnow collecting instruments, such as nets and bamboo poles as hazards, and 38% of the participants reported fish cutting instruments, such as knives, chopper (*da*) and traditional fish-knives (*boti)* as hazards causing injuries. 58% of the women suggested that they got injuries either due to fish sting or fish bites while handling the fishes for sorting, grading and processing for drying. 49% reported that just contact with some species of fish might cause allergic reactions and inflammations. Other physical hazards reported include sharp stones, corals or seashells submerged under water (2%), sharp edges of bamboo slices while making fishing basket (1%), grinding machine of *nappi* or dry fish paste (1%) and risk of drifting away while working near the coast, especially during collecting minnows (1%).

#### Physical hazards

Most women in the fisheries sector worked under open sky under direct sun as 99% of the participants reported they exposed to direct sunlight for more than 4 hours a day. 31% of the women reported that they got exposed to noise at their workplace, mostly from workplace chaos (29.5%), but also from engine rooms (1%), generators (0.5%) and *Nappi* grinding machine (0.5%).

#### Chemical hazards

Most of the women participated in the study reported that they got contact salt water (90%) and salt (81%) in their day-to-day work. They encountered salt mainly during dry fish processing or dry fish paste (nappi) processing. Some women (42%) reported to have contact with or exposed to chemical compounds, especially different kind of insecticides, during dry fish processing.

#### Ergonomic hazards

Majority of the women (91%) reported that in their occupation they need to work in uncomfortable body posture. 84% women had to sit in an uncomfortable position for long period. This happens especially during collecting fish from nets, collecting minnows from the coast, sorting and grading fishes, and cutting or processing fishes, when they have to sit on their feet without any support to hip or back. More than half of the participated women had to lift heavy weight, specially, fish and fish products.

#### Biological hazards

The study identified some biological hazards that may contribute to transmission of infectious diseases. Nearly three-fourth of the women reported that they did not provision of sanitary latrine (72.46%) and handwashing with soap (78.26%) at their workplaces. Nearly same proportion (74.88%) of the women reported that they need to remain wearing wet clothes for prolonged time.

Additionally, during FGDs, women from some fishing communities expressed that they did not have any specific time schedule for work or rest. They need to rush whenever the boat arrives. Parallel to the economic activities, the women in the fisher communities needed to maintain household chores and take care of their children. In some communities, even during off fishing season, the women got engaged in non-fisheries activities, such as working as house keepers.

### Occupational risks or health conditions

Table 4 summarized the occupational risks in terms of illnesses and health conditions resulting from the occupational hazards.

**Table 4.**
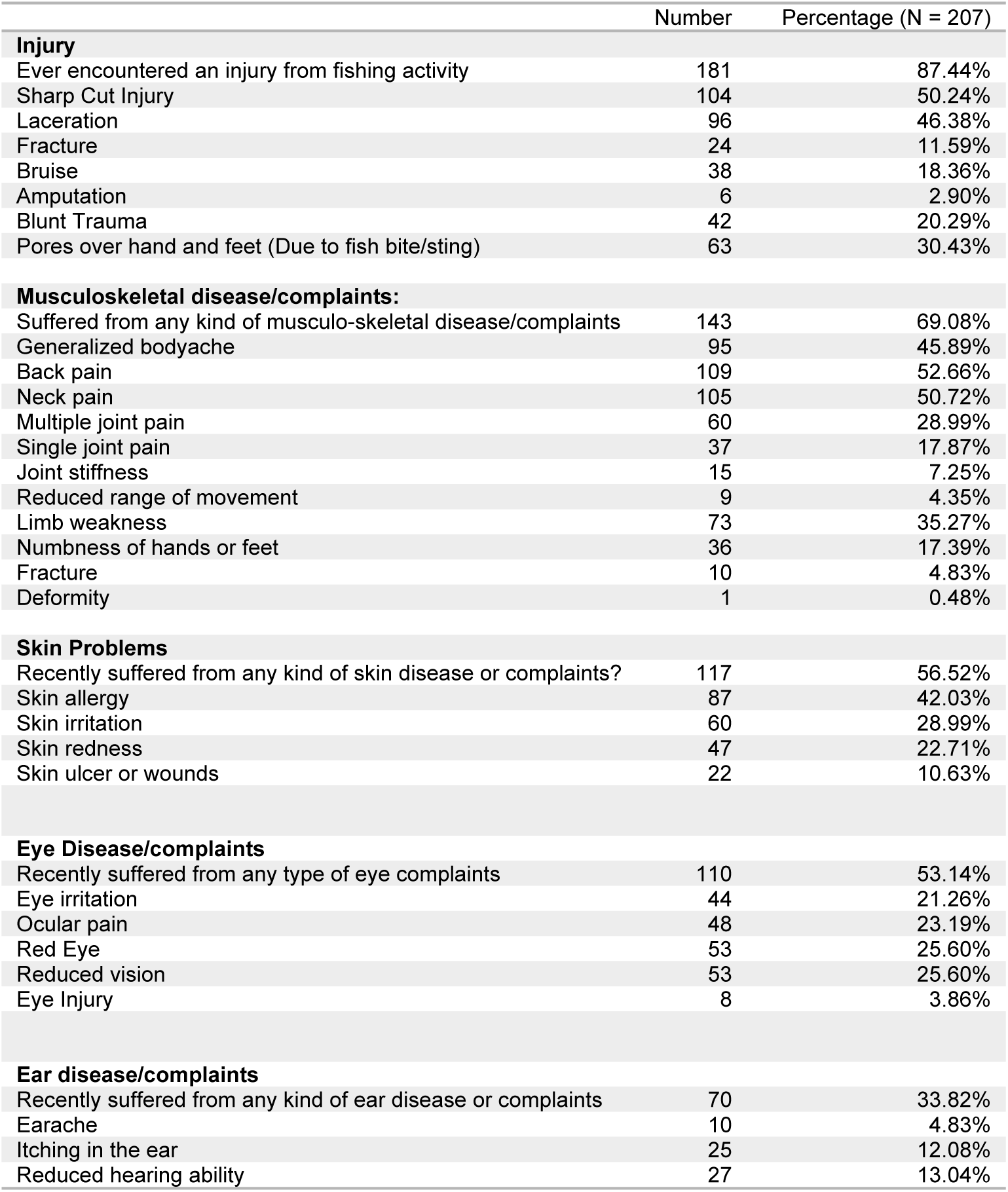
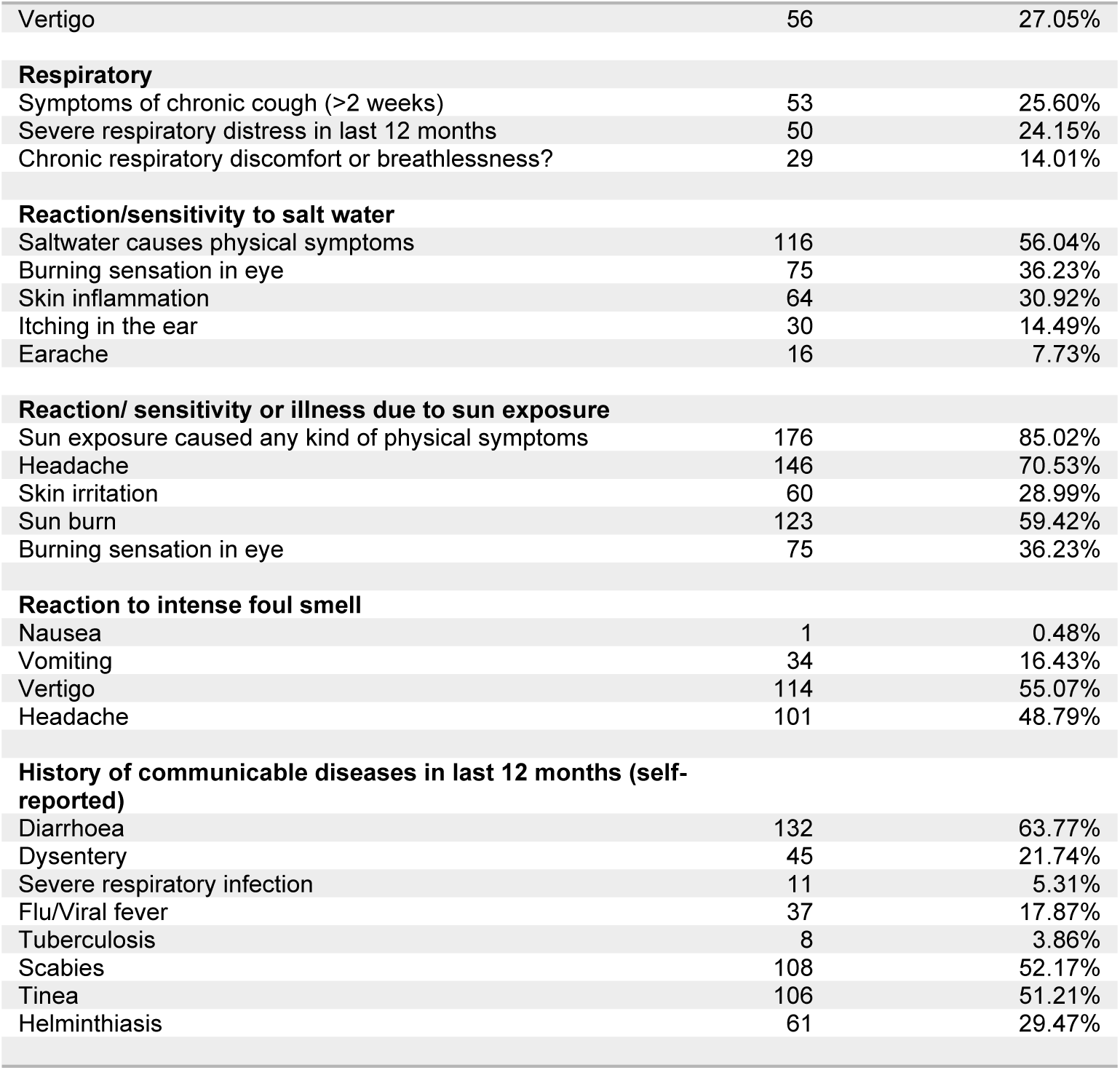
Occupational risks in terms of illnesses and health conditions.

#### Injuries

87.44% of the women engaged in fishery industry who participated in the study reported that they encountered an injury in their lifetime due to fishery activities. The injuries included sharp cut injury (50.24%), laceration (46.38%), fracture (11.59%), bruise (18.36%), amputation (2.90%) and blunt trauma injury (20.29%). Around one third of the participants (30.43%) reported that they developed multiple pores on their hand and feet due to repeated fish bites or stings. During focus group discussions, some women also expressed that few days after the injuries sometimes the wounds got infected.

#### Musculoskeletal disease/complaints

More than two-third of the women (69.08%) experienced a musculoskeletal disease condition, such as generalized body ache (45.89%), back pain (52.66%), neck pain (50.72%), multiple joint pain (28.99%), single joint pain (17.87%), joint stiffness (7.25%), reduced range of movement (4.35%), weakness of the limbs (35.27%), numbness of hands or feet (17.39%) and deformity (0.48%). Severity and chronicity of the disease varied among the participants – 24.64% reported as acute and severe, 33.82% reported acute but mild, 29.47% reported chronic and severe, and the rest 12.08% reported chronic mild.

#### Skin disease/symptoms

More than half of the respondents (56.5%) gave the history that they recently suffered from a skin disease or had a skin related symptom, which include skin allergy (42.03%), skin irritation (28.99%), redness of skin (22.71%), and ulceration or wounds over skin (10.63%).

#### Eye diseases/complaints

More than half of the respondents reported that they recently experienced eye related disease symptoms or had eye diseases, such as eye irritation (21.26%), ocular pain (23.19%), red eye (25.60%) and reduced vision (25.6%). 3.86% reported that they encountered an eye injury.

#### Ear disease/complaints

Around one-third of the responding women (33.82%) reported that they recently had an ear disease or experienced ear-related symptoms such as earache (4.83%), itching in the ear (12.08%), reduced hearing ability (13.04%), and vertigo (27.05%).

#### Respiratory illnesses/symptoms

A quarter of the participants reported that they had symptoms of chronic cough for more than two weeks. 24.15% reported that they had respiratory distress in last 12 months. 14.01% of the women complained that they had chronic respiratory discomfort or breathlessness.

#### Reaction/sensitivity to salt water

56.04% of the women participated in the study reported that exposure to saltwater caused them some kind of physical symptoms, which include burning sensation of eye (36.23%), skin irritation (30.92%), ear itching (14.49%) and earache (7.73%).

#### Reaction/ sensitivity or illness due to sun exposure

Majority of the respondents (85.02%) informed that prolonged exposure to sun ensued some physical complaints, including headache (70.53%), skin irritation (28.99%), and eye irritation (36.23%). 59.42% of the women experienced a sunburn.

#### Reaction to intense foul smell

Intense smell from fishes during dry fish or dry fish paste processing, or chemical compounds causes vertigo among 55.07%, headache among 48.79% and vomiting among 16.43% of the women.

#### History of communicable diseases in last 12 months

The self-reported incidence of communicable diseases was found very high among the women working in the fisheries sector. Communicable diseases experienced in last 12 months include diarrhoea (63.77%), dysentery (21.74%), severe respiratory infection (5.31%), flu/viral fever (17.87%), tuberculosis (3.86%), scabies (52.17%), tinea (51.21%), and helminthiasis (29.47%).

### Preventive and safety measures

Overall, 78.26% of the women responded that they did not use any personal protection equipment at their workplace. Table 5 summarized the use of different types of personal protection equipment by the women at their workplace.

**Table 5:** Use of personal protection equipment by the women working in fishery industry Use of PPE No Rarely Sometimes Often.

| Use of PPE | No | Rarely | Sometimes | Often |
| --- | --- | --- | --- | --- |
| Facemask | 143<br>(69.08%) | 37<br>(17.87%) | 26<br>(12.56%) | 1<br>(0.48%) |
| Hand gloves | 163<br>(78.74%) | 14<br>(6.76%) | 28<br>(13.53%) | 2<br>(0.97%) |
| Gumboot | 196<br>(94.69%) | 9<br>(4.35%) | 2<br>(0.97%) | 0<br>(0.00%) |
| Raincoat/similar rain protection | 124<br>(59.90%) | 25<br>(12.08%) | 34<br>(16.43%) | 13<br>(6.28%) |
| Sunglass | 202<br>(97.58%) | 4<br>(1.93%) | 1<br>(0.48%) | 0<br>(0.00%) |
| Sunscreen | 195<br>(94.20%) | 6<br>(2.90%) | 4<br>(1.93%) | 2<br>(0.97%) |

More than 69% of the women never used a mask in their workplace and 17.87% only used on rare occasions. 78.74% of the women never used any hand gloves in their workplace and 6.76% used it rarely. More than 94% of the women never used gumboot. More than 94% of the women never used sunscreen or sunglasses for sun protection. 59.90% of the women never used a raincoat or similar material for rain protection, 12.08% used it on rare occasion. As expressed by the women during FGDs, the common reasons for not using a PPE at workplace were unavailability of PPE, discouragement by the fish farm owners, and operational inconvenience.

93.72% of the women reported that they did not have any first aid kit at their workplace which could be accessed during an injury or emergency health condition.

Participants also expressed that the fishing farms did not take any liability for treatment or care in case the workers get injured or sick due to their occupation. There was no special measures or arrangement organized at the workplace for pregnant or lactating women. Payments were made based on daily activity, there was no system in place for maternity leave or benefits.

## Discussion

This study gives in-depth insights into the occupational hazards and health risks experienced by the women actively engaged in the fisheries sector in Cox’s Bazar and Chattogram. To the best of the author’s knowledge, this is the first study carried among the fisherwomen in Bangladesh revealing their occupational health hazards and risks. The study found a high occurrence of occupational hazards, health risks and disease conditions and limited availability of preventive measures among the women in the fisher communities in coastal areas of Bangladesh.

The demographic profile of our study participants reflects the distinctive characteristics of women working in the fisheries sector in Cox’s Bazar and Chattogram. We have found an alarming status of education among the women in fisheries sector, with most participants unable to sign or could merely sign, indicating high rate of illiteracy among the women in fisher communities. Literacy rate is comparatively very low among the fishermen in Bangladesh ^18–19^ and due to gender-based discrimination the rate is further low among women in the fisherfolk communities^5^. Such a lack of education may have an impact on their awareness of work hazards as well as their capacity to acquire and comprehend safety information and procedures. Therefore, awareness raising, and prevention strategies related to occupational hazards should be customized based on the education status of the women.

Our study revealed that women were engaged in mainly three types of fisheries industries - dry fish processing, raw fish processing and marketing and dry fish paste (*nappi*) processing, with majority of them (88.41%) dedicating direct manual labour. Traditionally, women in the coastal regions of Bangladesh are not engaged in fish harvesting, which is usually seen as occupation of man. This aligns with the social norms of majority of the world’s small scale fishing communities, where men are the primary producers and women get engaged in fish processing, marketing and distribution.^20^ Interestingly, a significant majority of women were involved in different fishing operations at the same time, demonstrating the variety and adaptability of their positions in this sector. Common daily chores were fish sorting and grading, fish cutting or scale removal, and minnow collection illustrating the multifarious nature of their employment. Women in the fisher communities also need to get engaged in household chores and caregiving to their children.^5^ Such double burden of household responsibilities and economic workload may have significant effect on their health.^21^

The study found a number of physical safety hazards that causes injuries to the women. Slippery surface in the workplace is a commonly reported hazard for women, which may not only cause acute injuries but also chronic musculoskeletal disorders. This finding is consistent with findings of other studies in Kenya^11^ and India^12^, which also reported accidental falls and injuries happening among fisher women due to the slippery surface. Our study found the high occurrence of injuries among the women working in fisheries sector, which include sharp cut injuries, laceration, fracture, bruise, blunt trauma and amputation. Injuries among fisherwomen were also evidenced in previous studies among fisherfolks of coastal states in India^22^, Irular tribal community^12^ and Fisherfolk in Kampi Samaki^11^. Although most of the injuries are minor and nonfatal, unable to properly treat the wound in a timely way may result in infection.^23^ In Cox’s Bazar and Chattogram coastal areas, as revealed by our study, the injuries can result from a range of workplace physical safety hazards such as fishing or minnow collecting instruments (e.g. nets), fish cutting instruments and fish stings or bites. Women engaged in dry fish processing need to cut or descale the fishes using sharp instruments, accidental handling of such instruments may result in the sharp cut injuries, laceration and even amputation of fingers.

Many of the women engage themselves in minnow collection at the seashores, specially, during monsoon season, using small fishing nets and bamboo poles. Some women reported cuts and trauma from such nets and poles. Although less common, a few women performing this activity also reported the risk of drifting away to sea while working near the coast. This finding could be consistent with the finding of Dutta^13^, who claimed women in Bangladesh engaged in fishing with their children possesses the risk of drowning since the traditional cloths are not flexible enough for free movement in the water. A few have reported injuries to their hands, legs, and feet from sharp stones, corals, or oysters submerged in water, particularly when working submerged in sea water for collecting minnow or prawns. This hazard for cut injury was also reported by Velvizhi and Gopalakrishnan^12^, who argued that wounds resulting from oyster shells could generally be very deep, bleed profusely and get infected.

More than half of the women identified fish sting or fish bite as an agent for their injuries. Most of the women working in the fisheries sector are to directly handle the fishes for sorting, grading, cutting, descaling or drying. Those who engaged in dry fish processing also need to make special knots near around gum of some fish species. Since they handle the fishes without any personal protection equipment (e.g. gloves), they commonly encounter the injuries from the fish spines or teeth. Often, such injury leaves with short- and long-term pores on hands and feet of the women. Such injuries caused by fish species is common among men and women working in the fishery sector and reported by large number of studies in Nigeria^8^, Kenya^11^ and India^12^. Fish stings and bites may also result in infections and subsequent amputation of fingers or toes.^24^

The study found high prevalence of musculoskeletal disease conditions among the women in fishery sector as reported by more than two-third of the interviewed women. This includes, generalized body ache, back pain, neck pain, joint pain and stiffness, reduced range of movement, muscle weakness, numbness of limbs and physical deformity. This is consistent with a study among fisherwomen in India, which found that 67% of the fisherwomen complaining of musculoskeletal pain and discomfort, especially for lower back, knee and upper back^22^. In our setting, this finding could be related to a wide range of ergonomic hazards identified by this study, such as working in uncomfortable body posture, prolonged sitting in an uncomfortable position and carrying heavy weight. One of the unique findings of this study is that we found women are to sit on their feet without any support to hip or back during their fish processing and minnow collecting activities. Such uncomfortable and unsupported body posture may result in different musculoskeletal conditions, including low back pain, cervical pain and joint pain. Women engaged in the fishery sector also need to manually carry heavy weight, such as fish and dry fish, during their fish processing activity. Heavy weight carrying as a factor for musculoskeletal disease conditions among fisherwomen was also reported in previous studies^11^. Many of the women also need walk long distance for collecting and delivery of fish and fish products in our setting, which also may lead to musculoskeletal pain. This is consistent with the finding of the study among the Irular fisherwomen.^12^

Although, use of pesticide in dry fish industry is strictly prohibited by the law in Bangladesh, some women informed that they get exposed to pesticides in their dry fish farm. Occupational exposure of pesticide can be associated with respiratory health problems, including asthma, chronic obstructive pulmonary disease (COPD) and lung cancer. ^25^ Establishing an association between exposures of the fisherwomen to pesticide and developing different clinical conditions was beyond the scope of this research, since it requires clinical diagnosis of the cases. However, our study found a high rate of occurrence of self-reported respiratory complaints, especially chronic cough, respiratory distress and chronic respiratory discomfort/breathlessness among the fisherwomen. Further research is warranted to explore the association between chemical exposure and respiratory diseases.

Our study found that majority of the women get frequent contact with salt water (90%) and salt (81%) in their daily activities, and more than half of the women reported that they develop some kind of physical symptoms, such as eye irritation, skin irritation, ear itching and earache, in reaction to the salt water. We have also noted a high incidence of skin diseases among the women in fisher communities, including skin allergy, irritation, ulceration and wounds. Salt or salt water could be the potential factor for such incidents as a recent study^26^ found that sodium chloride could be a potential factor in the progression of allergy in the skin. This could also be related to contact with certain fish species that provoke allergic reactions.^27^ Velvizhi and Gopalakrishnan^12^ argued that exposure to brackish water, jellyfish and algal bloom can result in skin irritation, rashes and psoriasis among fisherwomen. However, further research is warranted to statistically establish the assumption.

The study found that nearly all (99%) of the women got exposed to direct sunlight for more than 4 hours a day and majority of them reported some kind of reaction or sensitivity to sun exposure, which included headache, skin irritation, sun burn and eye irritation. Ultraviolet radiation of sunray may result in acute sunburn and chronic skin changes, including skin cancer.^28^ Sunburn among fisherwomen was also reported as a health concern in Kenya.^11^ Kyei et al.^29^ identified seawater and sunrays as ocular hazards causing ocular irritation, tearing, red eyes and blurred vision. We also found high incidence of eye related diseases or symptoms among the women in fisherfolk, which include eye irritation, ocular pain, red eye and reduced vision. These could be related to the prolonged exposure to sun and/or saltwater.

Fisher folks in Chattogram and Cox’s Bazar live in overcrowded settings with fragile shelter and compromised water, sanitation and hygienic practices. Our study found that more than three-fourth of the women in the fisherfolk did not have the provision of sanitary latrine and handwashing with soap at their workplaces. All these factors can contribute to a high burden of communicable diseases among the fisherfolk community. This could be evidenced by the fact, as found in our study, of a very high incidence of self-reported communicable diseases, specially, diarrhea, dysentery, flue/viral fever, respiratory infection, tuberculosis, skin infections and helminthiasis among the women in fisher communities. Moreover, we also found that nearly three-fourth of the women needed to remain wearing wet clothes for prolonged time. Weather in Bangladesh is hot and humid, especially during the summer. Both these factors – wet clothing and warm weather are risk factors for tinea^30^. Concurring this, in our study we found that half of the women had self-reported incidence of tinea in last 12 months. Although, the reported incidents communicable diseases were not clinically verified, intensified health promotion and risk communication efforts are recommended both at community level and at workplace to improve the hygienic behavior and practices.

Many of the hazards could be mitigated by use of personal protective equipment, such as gloves, masks, sunglasses, sunscreen and gumboots.^14^ However, we have found majority of the women in the fisher communities did not use any personal protection equipment at their workplace. While use of gloves could prevent many of the injuries associated with fish bite or sting and sharp instruments, commonly available heavy-duty gloves may not be feasible for fisherwomen since these are not suitable to use for fine work carried out by the women during fish processing. Therefore, personal protection equipment should be innovated or customized considering the local need, context and requirement. Moreover, the fisherwomen should be trained and mobilized for using personal protection equipment for their health and safety. Finally, we have found that the first aid kits were grossly unavailable at the workplace, with which injuries could be managed early and risk of wound infection could be prevented. Strong advocacy is needed to influence the fish industry players to make personal protection equipment and first aid kits available at the workplace.

### Limitations of the Study

The study reported the hazards and health conditions based on the experiences shared by the participants. The conditions were not clinically validated. However, the author tried to mitigate the response bias by triangulation of qualitative and quantitative data and appropriate training of the enumerators on case definitions of clinical conditions. The study was also only descriptive in nature and did not have the component of analyzing the clinical correlation. Another limitation of the study was it did not include mental health and psychosocial aspects of the participants.

## Conclusion and Recommendations

Women in the fisher communities in Bangladesh are engaged in a wide range of activities in the fisheries sector. Our study found a high occurrence of occupational hazards experienced by the women, including physical safety, physical, chemical, ergonomic and biologic hazards. Due to unavailability of adequate preventive and safety measures, the hazards are resulting in a wide range of health risks and disease conditions among the women in the fisher communities, including injuries, musculoskeletal disorders, skin diseases, eye diseases, respiratory diseases and communicable diseases. However, their workplaces are pervasively missing with the availability of personal protection equipment, first aid kits and health and safety benefits. Notwithstanding the adversities, the women continue to work hard in the fishing industry, even though their efforts are merely acknowledged in the society. Therefore, a comprehensive multisectoral strategy, considering the below recommendations, needs to be undertaken to mitigate the occupational hazards and prevent associated diseases among the women in fisher communities promoting their health and wellbeing.

1. The women working in the fisheries sector should be trained on the occupational hazards and risks, and safety and preventive measures.
2. Procedure should be innovated or identified in consultation with fisheries sector experts regarding safe handling of fish and associated instruments to prevent injuries from physical safety hazards.
3. First aid kits should be made available at all workplaces and a focal person from the workers should be trained in the use of the kits.
4. Engineering and ergonomic innovations are necessary to prevent ergonomic hazards. This could include proper sitting arrangement, measures to avoid prolonged standing and convenient tools (e.g. carts, wheel barrows) to carry heavy weight.
5. Personal protection equipment should be made available to the women in the fisher folk and train and motivate them on their use. The PPE should be customized for use in the fisheries sector context.
6. Advocacy should be conducted to orient the fish industry owners on the occupational hazards, so that they improve the workplace safety and ergonomics and arrange measures to prevent physical hazards, including direct sun exposure and noise, and ensure use and supply of personal protection equipment.
7. Hygienic conditions should be improved at both workplace and personal level, including arrangement of sanitary latrines, soap and water for handwashing, safe drinking water and promotion of personal hygiene.
8. Restrict use of insecticide in dry fish processing and promote organic method of dry fish processing to prevent exposure to chemical hazards.
9. Establish a mechanism for regular health checkup of the women engaged in the fisheries sector to prevent disease or illness at early stage.
10. Advocacy should be conducted with the owner of the fisher industries to ensure maternal benefits, menstrual hygiene mechanisms at workplace.

## Data Availability

All relevant data are within the manuscript and its Supporting Information files.

## Acknowledgement

The authors acknowledge Bright Bangladesh Forum and its staff, specially, Utpal Barua and Mahfuzur Rahman for arranging operational support for this study. We also thankful to the enumerators of the study, Tajmin, Abdullah, Mahbuba and Sawrin, for their support in quantitative data collection and field coordination.

## Supporting information

**S1. Dataset of the study.** Dataset of the study is attached as reference.

